# Contact tracing strategies for COVID-19 containment with attenuated physical distancing

**DOI:** 10.1101/2020.05.05.20091280

**Authors:** Alyssa Bilinski, Farzad Mostashari, Joshua A Salomon

## Abstract

Contact tracing has been recommended as a critical component of containment strategies for COVID-19. We used a simple epidemic model to evaluate how contact tracing might enable modification of current physical distancing restrictions. Testing and tracing coverage need to exceed 50% to see substantial gains; if both are below 50%, contact tracing does not reduce transmission by more than 10%. With 90% testing and tracing as well as high isolation and quarantine efficacy, contact tracing could reduce overall transmission by >45%, which would allow for partial loosening of physical distancing measures. Benefits of contact tracing could be enhanced by testing all contacts rather than only those with symptoms and by policies to support high adherence to voluntary isolation and quarantine.

## Introduction

As of May 1, the novel coronavirus SARS-CoV-2 has infected more than 3.2 million people worldwide and caused over 233,000 deaths.^1^ Since March, most communities in the United States have been living under physical distancing measures including stay-at-home orders in 40 states.^2^ Evidence suggests that these mitigation efforts have slowed the spread of the virus in many jurisdictions, and as of mid-May, more than 30 states have moved to loosen these measures.^3,4^ A number of frameworks have been proposed for the safe relaxation of non-pharmaceutical interventions, and most include scaling up testing and contact tracing to support containment.^5-9^

Guidelines for contact tracing call for identifying and monitoring individuals who have been in close contact with confirmed positive cases, facilitation of testing for symptomatic contacts, and counseling and follow-up to encourage voluntary self-isolation, quarantine and symptom monitoring.^10,11^ Several previous papers have considered the role of contact tracing for containment of COVID-19,^12-15^ but important questions remain about potential impact given uncertainty around the extent of presymptomatic and asymptomatic transmission of SARS-CoV-2 and the efficacy of voluntary isolation and quarantine. As decision-makers look toward relaxing current physical distancing measures, there is an urgent need to quantify the degree to which contact tracing programs could allow for partial loosening of restrictions while maintaining control over resurgent infection.

This paper uses a simple model to evaluate different contact tracing strategies to support modification of physical distancing restrictions. We examine the necessary conditions for maximizing benefits of contact tracing. We consider how broadening current testing guidelines from the Centers for Disease Control and Prevention to include testing for contacts without symptoms could amplify the impact of contact tracing programs.

## Methods

We developed a simple deterministic Markov branching model of COVID-19 (**Figure S1**). Epidemiological parameters were adapted from prior modeling studies where available (**Table S1**). Infected individuals generate new infections based on whether they have symptoms, whether disease is detected, and whether they have been identified as a contact of an infected individual (**Table S2**). In our base case analysis we assumed that 40% of infections are asymptomatic (varied to 20% in sensitivity analysis),^16-19^ and that confirmed cases have 50% lower rates of transmission than unconfirmed cases. We assumed that symptomatic cases become infectious prior to emergence of symptoms.^20,21^

Estimates on the effectiveness of contact tracing vary considerably.^22,23^ We modeled an array of different scenarios in order to characterize prerequisites for effective contact tracing, as well as to evaluate different possible policy priorities. We defined scenarios by the fraction of symptomatic cases detected in the community (not linked to a tracked case), the fraction of contacts successfully traced, the isolation and quarantine efficacy among traced but undetected contacts, and whether testing was restricted to those with symptoms or includes all traced contacts (**Table 1**). Given the likely importance of levels of community testing as a prerequisite condition for contact tracing, we conducted a secondary analysis that quantified the combined benefit of scaling up both testing and contact tracing against a counterfactual in which detection of symptomatic cases remains constant at an assumed current fraction of 20% and detection of asymptomatic cases at 5%.

**Table 1.** Parameters defining contract tracing scenarios.

| Parameter | Range |
| --- | --- |
| Fraction of symptomatic cases detected | 10-90% |
| Fraction of contacts successfully traced | 10-90% |
| Isolation and quarantine efficacy<br>(% reduction in transmission) | 30%, 60%, 90% |
| Testing strategy | Test symptomatic only or<br>test all contacts |

We evaluated the impact of different contact tracing strategies in terms of the percentage reduction in the effective reproductive number *R_t_* (average number of secondary infections produced by each infection) under each contact tracing scenario, compared to a scenario without contact tracing. Assuming that contact tracing strategies would be implemented alongside policy changes to partially relax physical distancing measures, and that the containment phase would begin when *R_t_* was less than or equal to 1.0, reductions in *R_t_* can be used to compute the *containment margin* for a given strategy. The containment margin signals how much current physical distancing measures could be relaxed in the presence of contact tracing, while maintaining *R_t_* below the critical threshold of 1.0.

## Results

### Base case

Base-case results are summarized in **Figure 1** (see **Figure S4** for details). Both community detection of symptomatic cases that are not linked to a tracked case and successful tracing of contacts needed to be at least 50% to see substantial gains; when both were less than 50%, no contact tracing strategy reduced *R_t_* by more than 10% compared to corresponding scenarios without contact tracing.

**Figure 1.**
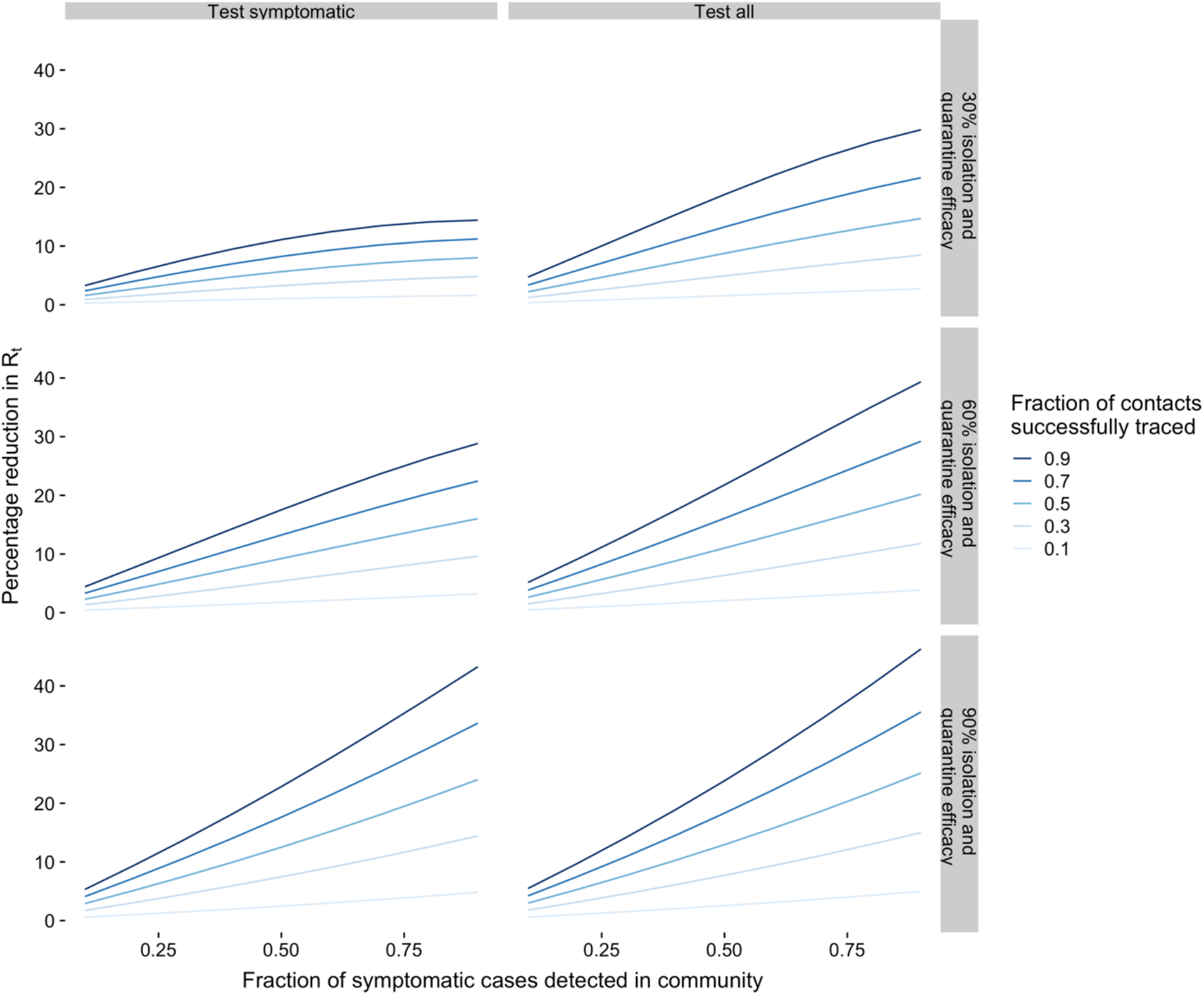
Effects of contact tracing strategies under varying assumptions about key program features. The horizontal axis shows the fraction of symptomatic cases that are detected in the community. The vertical axis shows the primary measure of strategy effectiveness: the percentage reduction in *R_t_* in the contact tracing scenario relative to *R_t_* without contract tracing. The color of the lines within each panel indicate the fraction of contacts that are successfully traced. ‘Isolation and quarantine efficacy’ refers to the level of reduction in transmission rates from traced, undetected contacts.

Testing asymptomatic contacts may substantially increase the impact of contact tracing strategies. Across all scenarios with adequate fractions (>50%) of symptomatic cases detected in the community and contacts traced, testing asymptomatic contacts increased the benefit of contact tracing by a median factor of 1.3 (IQR: 1.1 to 1.7). Benefits of asymptomatic testing were larger when isolation and quarantine was less than 90%.

The overall impact of contact tracing depends strongly on isolation and quarantine efficacy. Median reductions in *R_t_* assuming isolation and quarantine efficacy of 30%, 60% or 90% were 11%, 21% and 29% respectively, for strategies that tested only symptomatic contacts, and 21%, 26% and 30% for strategies that tested all contacts. The contact tracing scenario with the greatest impact overall–defined by high levels of symptomatic detection and successful tracing, high isolation and quarantine efficacy, and testing of all contacts irrespective of symptoms—reduced *R_t_* by 46%.

### Sensitivity analysis

In a sensitivity analysis (**Figures S2/S5**), we considered how the potential impact of contact tracing strategies might vary if the percentage of cases without symptoms were 20%, rather than 40%. This increased effectiveness for all scenarios, by a median factor of 1.27 (IQR: 1.22 to 1.3), with a maximum reduction of 59%. We also evaluated the combined effect of scaling up both testing and contact tracing against the counterfactual of persistently limited testing at 20% of symptomatic cases and 5% of symptomatic cases and no contact tracing (**Figures S3/S6**). Accounting for both expanded testing and contact tracing together, the maximum reduction in *R_t_* increased to 57%, and the benefits in many scenarios were at least 10 percentage points greater than the benefits of contact tracing alone.

### Containment margins

To translate results into implications for potential modification of current policies, we used the percentage reductions in *R_t_* from each contact tracing scenario to compute a corresponding containment margin, which indicates how much current physical distancing measures could be relaxed with contact tracing in place, while holding *R_t_* below 1.0. As an example, assume that current physical distancing measures have reduced the reproductive number from *R_0_* = 2.4 to *R_t_* = 1.0, and that a contact tracing strategy could reduce *R_t_* by 40%. Under these parameters, containment would be possible if relaxed physical distancing measures on their own could maintain *R_t_* below 1.67, because the further reduction by a factor of 0.6 due to contact tracing would bring *R_t_* below 1.0. This implies that together with contact tracing, physical distancing measures could be applied at 52% of their current, full implementation effectiveness and still maintain the critical containment threshold of R_t_<1.

If *R_t_* has been reduced to levels well below 1.0, the containment margin is greater (i.e. physical distancing measures could be further relaxed); if contact strategies are less effective the margin for loosening physical distancing shrinks. For example, if a contact tracing strategy were half as effective, producing a reduction in *R_t_* of 20%, physical distancing measures could only be reduced to 82% of their current intensity. Further examples of containment margins under different assumptions about *R_0_* and the benefits of contact tracing are provided in **Table 2**.

**Table 2.** Containment margin under different assumptions about R_0_ and the percentage reduction in *R_t_* from contact tracing. We assume that R_t_=1 before contact tracing is introduced. The containment margin signals how much current physical distancing measures could be relaxed in the presence of contact tracing, while maintaining *R_t_* below the critical threshold of 1.0. For example, if the containment margin is 82%, then physical distancing measures could safely be reduced to 82% of their current intensity after contact tracing is introduced. Lower containment margins imply greater room to relax restrictions.

| $R_0$ | Percentage reduction in $R_t$ from contact tracing | Containment margin |
| --- | --- | --- |
| 2.4 | 20% | 82% |
| 2.4 | 40% | 52% |
| 2.4 | 55% | 13% |
| 3 | 20% | 88% |
| 3 | 40% | 67% |
| 3 | 55% | 39% |

## Discussion

In this study, we computed expected reductions in the effective reproductive number, *R_t_*, under different contact tracing scenarios to quantify the degree to which contact tracing can allow for modification of public health orders and physical distancing restrictions while maintaining containment. To support containment, contact tracing must be implemented in concert with wide-scale community testing and must successfully track a high fraction of infected contacts. Our results indicate that contact tracing will support a partial relaxation of physical distancing measures but not a complete return to levels of contact prior to physical distancing, consistent with prior studies.^14,15^ For example, a recent paper estimated that adding contact tracing to self-isolation could reduce *R_t_* by 35-47%, assuming 90% compliance,^14^ which is similar to the ranges estimated in our analysis.

Testing of asymptomatic contacts could magnify potential benefits by extending the coverage of tracing and potentially contributing to improved efficacy of isolation and quarantine. Another potential benefit of testing asymptomatic contacts, not captured in our model, is that negative test results could reduce the number of people needing to quarantine presumptively or could reduce the duration of quarantine, which might produce health and economic benefits.^24^ The benefits of contact tracing also depend substantially on levels of adherence to isolation and quarantine among traced cases, which could be enhanced through policies such as providing voluntary out-of-home accommodations and income replacement.

Limitations of this analysis include a simplified modeling framework that lacks network or household structure, and also does not explicitly capture nursing homes, workplaces, or other potentially high-transmission venues. Many uncertainties persist, including the extent of asymptomatic prevalence and transmission. Nevertheless, by examining a range of scenarios that reflect key sources of uncertainty and policy-relevant variables, we provide benchmarks that can aid in developing evidence-based containment strategies to minimize the risk of resurgent COVID-19 spread.

## Data Availability

No data used

## Supplemental Information

**Figure S1.**
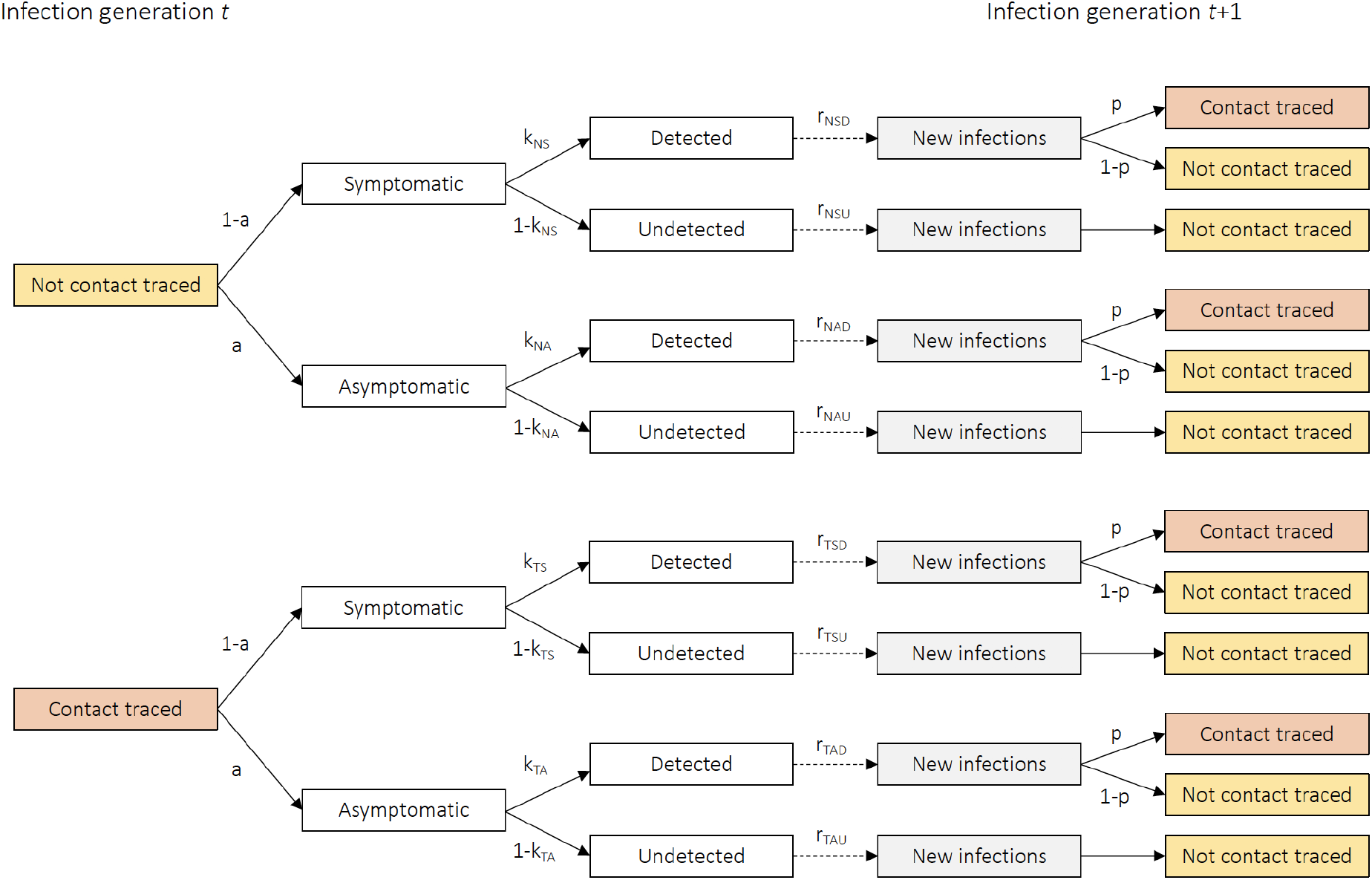
Model structure and parameters. Parameter definitions: *a* is the fraction of infections that are asymptomatic; *k* is the fraction of infections that are detected; *r* is the number of secondary infections from each infection and *p* is the fraction of cases that are successfully contact traced. For parameters indexed by subscripts: *T* is contact traced, *N* is not contact traced; *S* is symptomatic, *A* is asymptomatic; *D* is detected, *U* is undetected.

**Table S1.** Model parameter values.

| Parameter | Value | Notes |
| --- | --- | --- |
| Fraction of infections that are asymptomatic ($a$) | 40% | Estimates vary across studies.<sup>16–19</sup> Alternative value of 20% examined in sensitivity analysis (**Figure S2**). |
| Fraction of cases detected |  |  |
| Symptomatic, not contact traced ($k_{NS}$) | varies | Values shown in **Table 1** |
| Asymptomatic, not contact traced ($k_{NA}$) | 10% | Assumed to be small based on current US testing guidelines but that it will increase as key populations are screened<sup>25</sup> |
| Symptomatic, contact traced ($k_{TS}$) | 90% | Assumption, reflecting referral to testing for traced contacts |
| Asymptomatic, contact traced ($k_{TA}$) | 90% | Assumption, reflecting referral to testing. Applies only in contact tracing strategies that include testing for asymptomatics (see **Table 1**). |
| Number of secondary infections from each infection ($r$) | computed | See **Table S2** for details. |
| Fraction of cases successfully traced ($p$) | varies | Values shown in **Table 1**. |
| Duration of infectiousness |  |  |
| Presymptomatic ($d_P$) | 1.5 days | Durations inferred from temporal dynamics of viral shedding.<sup>20</sup> |
| Symptomatic ($d_S$) | 4 days | |
| Asymptomatic ($d_A$) | 5.5 days | |
| Relative infectiousness of asymptomatic infection compared to symptomatic infection ($v_A$) | 0.7 | Estimates vary across studies (e.g. 1.0,<sup>20</sup> 0.66,<sup>21</sup> 0.5,<sup>4</sup> 0.25<sup>26</sup>) |
| Relative infectiousness of presymptomatic infection compared to symptomatic infection ($v_P$) | 1 | Inferred from studies indicating substantial presymptomatic transmission.<sup>20, 21</sup> |
| Relative number of secondary infections from detected infections compared to undetected infections ($q$) | 0.5 | Limited empirical data, rationale for reduced secondary transmission includes: potentially increased likelihood of adherence to self-isolation, targeting of confirmed cases for public health support.<sup>27</sup> Alternative value of 1.0 examined in sensitivity analysis (**Figure** |
|  |  | **S3).** |
| Average daily rate of transmission for symptomatic cases not traced (*b*) | Calibrated | Values calibrated to produce baseline $R_t=1$. Note that relative reductions in secondary infections across program comparisons are scale invariant. |
| Isolation and quarantine efficacy (*e*) | Varies | Values shown in **Table 1**. Isolation and quarantine efficacy is approximately the product of how much infectious time remains when the contact is notified, and the degree of adherence to isolation and quarantine measures. Estimates of adherence have ranged considerably in previous studies (0-94%)<sup>22</sup>, including 70%<sup>21</sup> and 90%<sup>14</sup> in previous COVID-19 analyses. Remaining infectious time is difficult to measure, but likely less than 1.<sup>14,15</sup> A prior modeling study used efficacy estimates of 25% for a 'low-feasibility setting' and 75% for a 'high-feasibility setting.'<sup>12</sup> |

**Table S2.** Estimation of secondary infections.

| Category | Formula |
| --- | --- |
| Not contact traced, symptomatic, detected | $r_{NSD} = bd_P v_P + bd_S q$ |
| Not contact traced, symptomatic, undetected | $r_{NSU} = bd_P v_P + bd_S$ |
| Not contact traced, asymptomatic, detected | $r_{NAD} = bv_A d_A q$ |
| Not contact traced, asymptomatic, undetected | $r_{NAU} = bv_A d_A$ |
| Contact traced, symptomatic, detected | $r_{TSD} = (1-e)bd_P v_P + (1-e)bd_S q$ |
| Contact traced, symptomatic, undetected | $r_{TSU} = (1-e)bd_P v_P + (1-e)bd_S$ |
| Contact traced, asymptomatic, detected | $r_{TAD} = (1-e)bv_A d_A q$ |
| Contact traced, asymptomatic, undetected | $r_{TAU} = (1-e)bv_A d_A$ |

**Figure S2.**
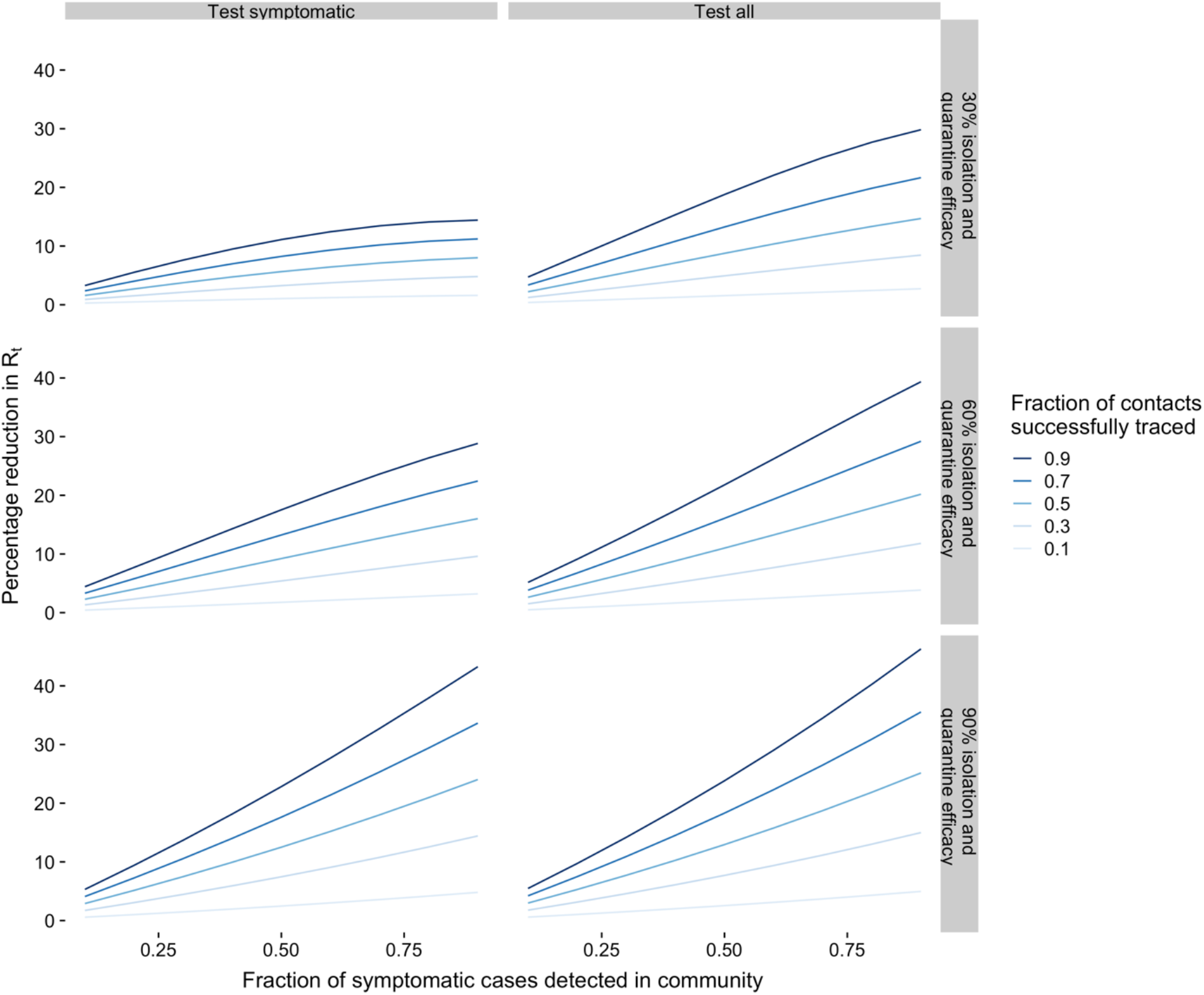
Results from sensitivity analysis assuming 20% of cases are asymptomatic. The horizontal axis shows the fraction of symptomatic cases that are detected in the community. The vertical axis shows the primary measure of strategy effectiveness: the percentage reduction in *R_t_* in the contact tracing scenario relative to *R_t_* without contract tracing. The color of the lines within each panel indicate the fraction of contacts that are successfully traced. ‘Isolation and quarantine efficacy’ refers to the level of reduction in transmission rates from traced, undetected contacts.

**Figure S3.**
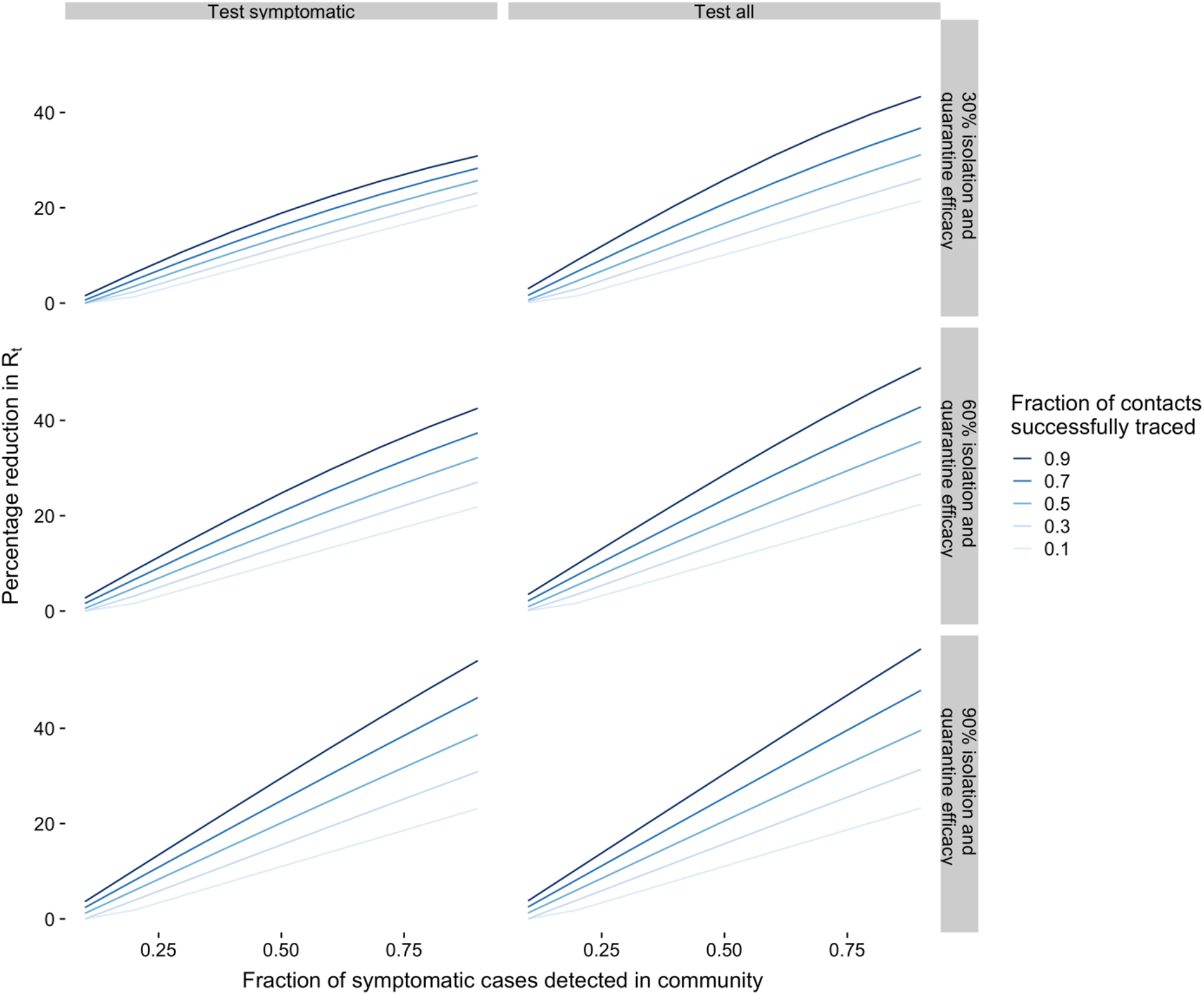
Results from secondary analysis on combined effects of scaling testing and contact tracing. The horizontal axis shows the fraction of symptomatic cases that are detected in the community. The vertical axis shows the primary measure of strategy effectiveness: the percentage reduction in *R_t_* in the increased testing plus contact tracing scenario relative to *R_t_* without increased testing or contract tracing. The color of the lines within each panel indicate the fraction of contacts that are successfully traced. ‘Isolation and quarantine efficacy’ refers to the level of reduction in transmission rates from traced, undetected contacts.

**Figure S4.**
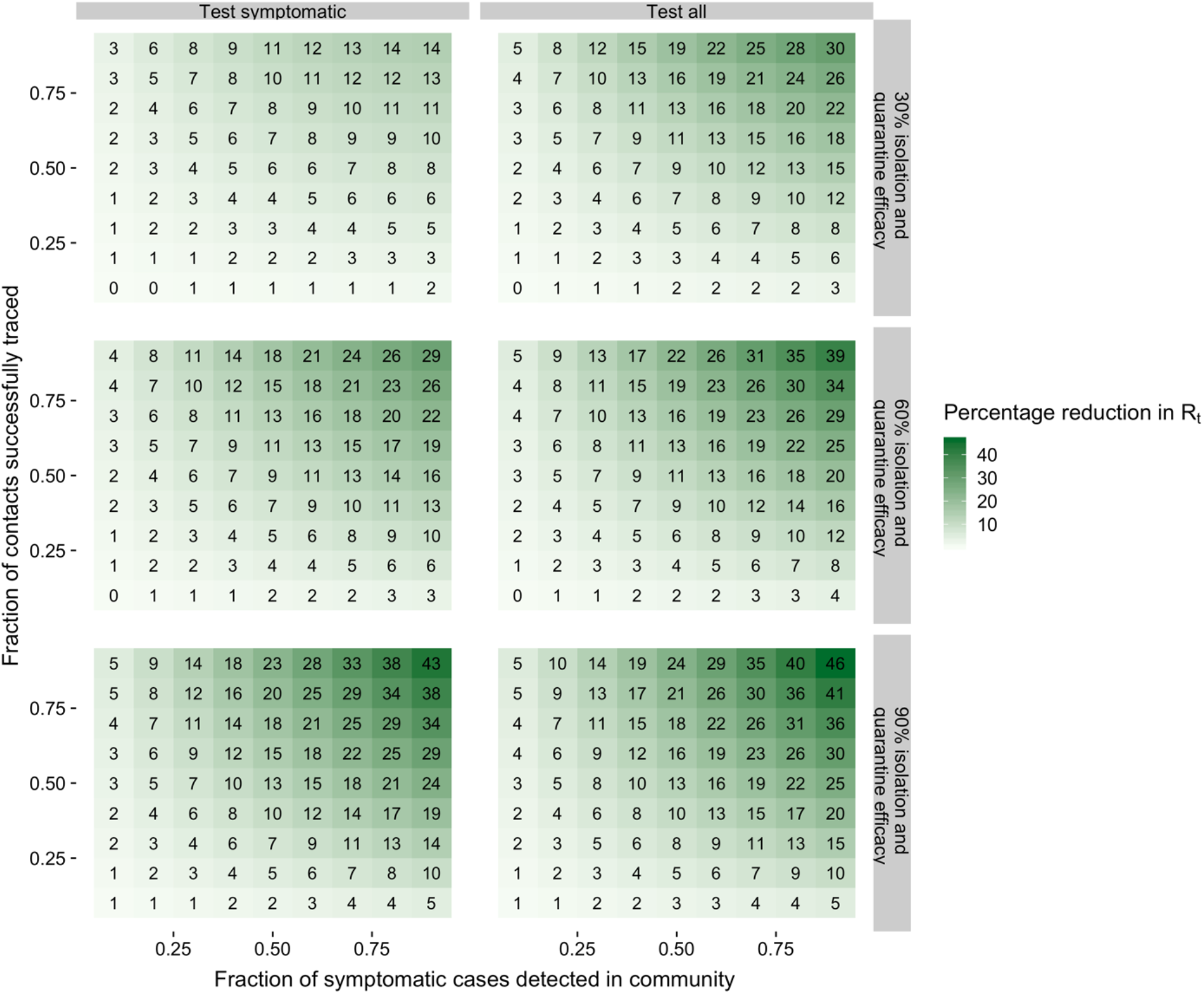
Heatmap representation of Figure 1. The horizontal axis shows the fraction of symptomatic cases that are detected in the community. The vertical axis shows the contacts that are successfully traced. The values and shading in each cell indicate the percentage reduction in R_t_ in the contact tracing scenario relative to R_t_ with no contract tracing. ‘Isolation and quarantine efficacy’ refers to the level of reduction in transmission among traced, undetected contacts.

**Figure S5.**
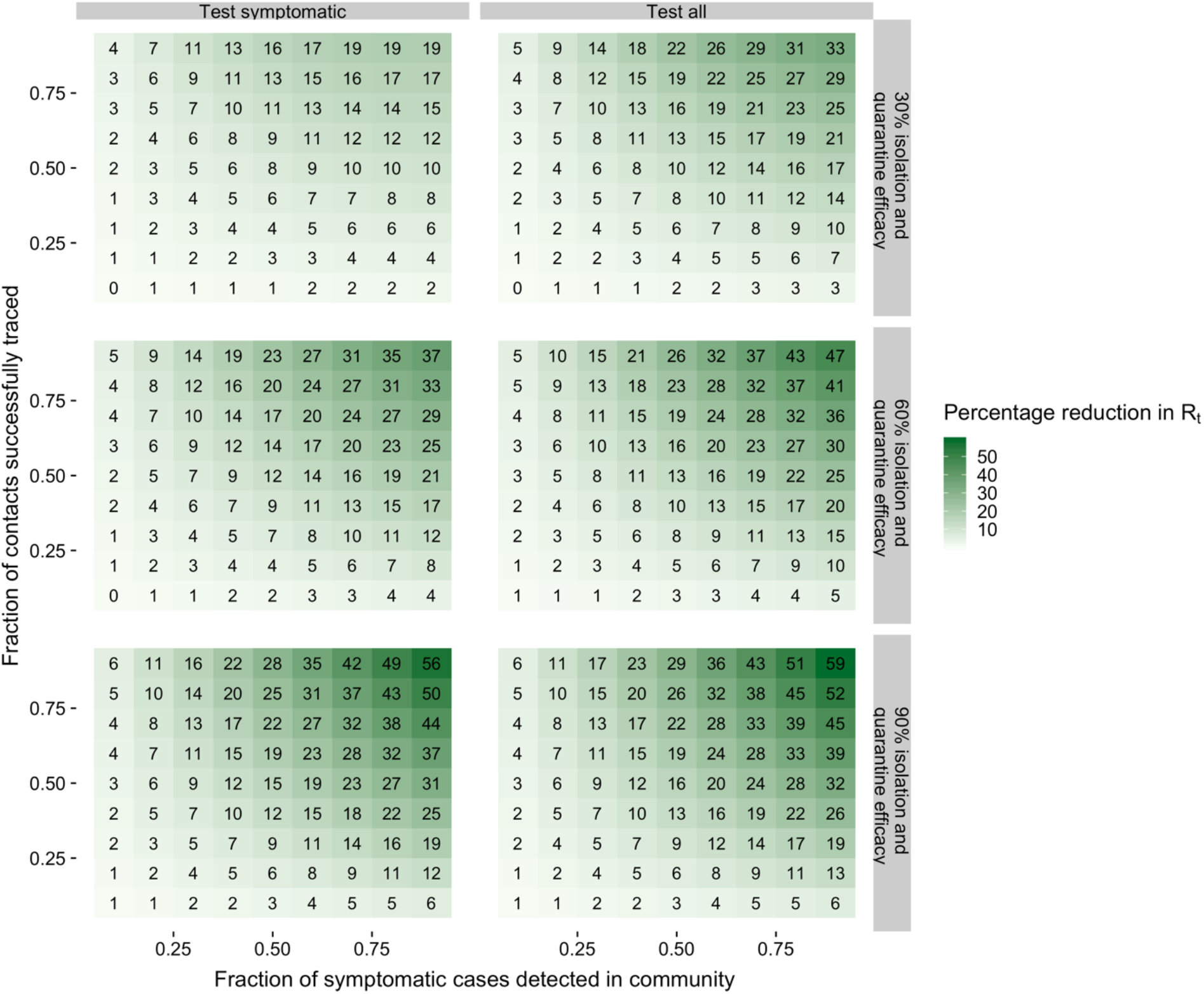
Heatmap representation of Figure S2: sensitivity analysis assuming 20% of cases are asymptomatic. The horizontal axis shows the fraction of symptomatic cases that are detected in the community. The vertical axis shows the contacts that are successfully traced. The values and shading in each cell indicate the percentage reduction in R_t_ in the contact tracing scenario relative to R_t_ with no contract tracing. ‘Isolation and quarantine efficacy’ refers to the level of reduction in transmission among traced, undetected contacts.

**Figure S6.**
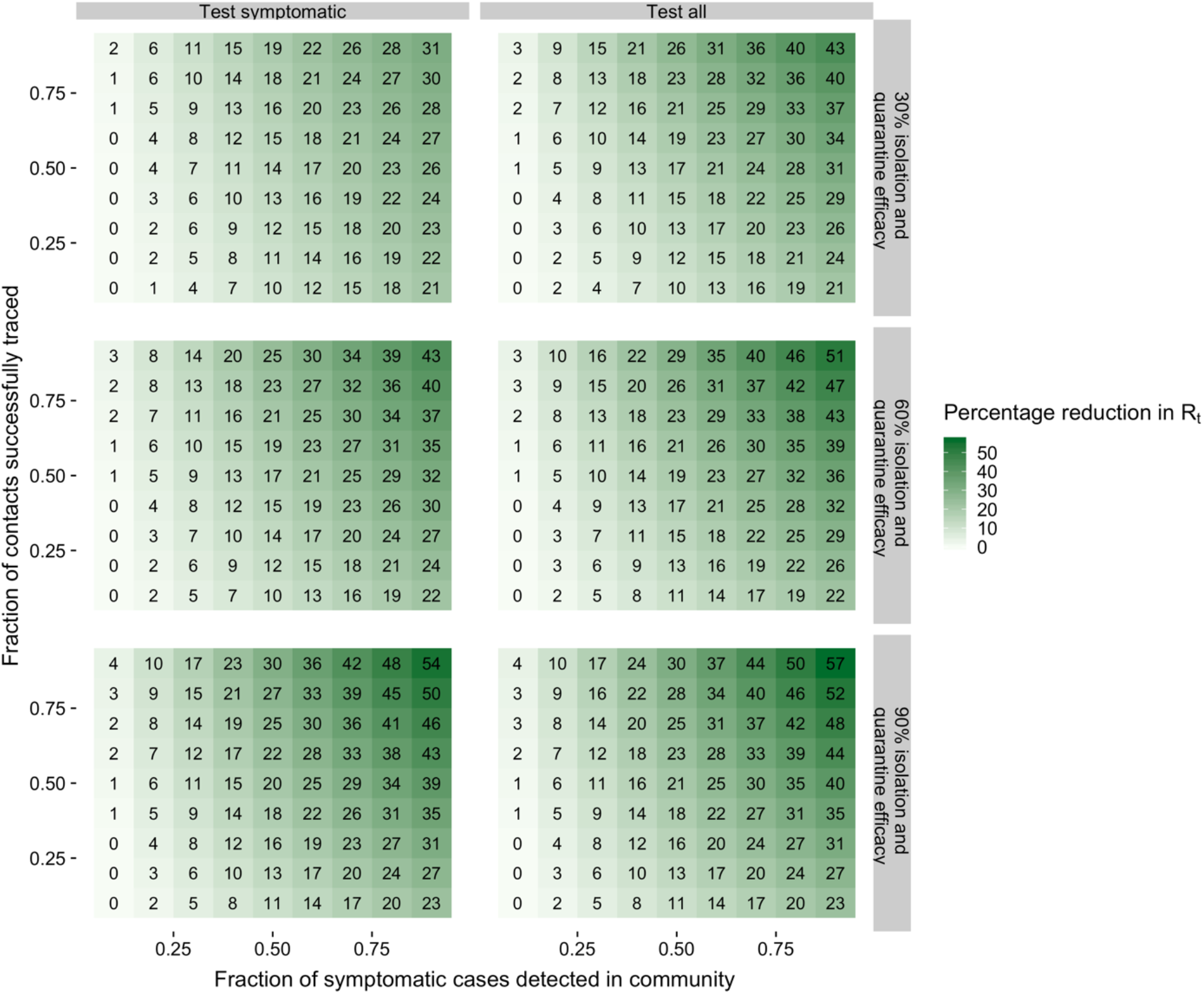
Heatmap representation of Figure S3: secondary analysis on combined effects of scaling testing and contact tracing. The horizontal axis shows the fraction of symptomatic cases that are detected in the community. The vertical axis shows the contacts that are successfully traced. The values and shading in each cell indicate the percentage reduction in *R_t_* in the increased testing plus contact tracing scenario relative to *R_t_* without increased testing or contract tracing. ‘Isolation and quarantine efficacy’ refers to the level of reduction in transmission among traced, undetected contacts.

